# Patterns of engagement in care during clients’ first 12 months after HIV treatment initiation in Zambia: a retrospective cohort analysis using routinely collected data

**DOI:** 10.1101/2024.10.03.24314849

**Authors:** Mariet Benade, Mhairi Maskew, Philip Chilembo, Mwanza Wa Mwanza, Theodora Savory, Brooke E Nichols, Carolyn Bolton Moore, Lloyd Mulenga, Suilanji Sivile, Khozya Zyambo, Sydney Rosen

## Abstract

**Background:** The first year after HIV treatment initiation or re-initiation is the period of highest risk of a treatment interruption or disengagement, yet little is known about the timing, patterns, and effects of interruptions in the early treatment period.

**Methods:** Using routinely collected electronic medical record data from 543 Zambian facilities from 01/01/2018 to 28/02/2023, we described patterns of engagement during the first year of HIV treatment. We categorized clinic visits and other interactions based on whether they were attended as planned (≤scheduled date), late ≤28 days, or late >28 days). We used these visit categories to define engagement patterns for months 0-6 and months 7-12 after initiation or re-initiation as 1) continuous (attended all scheduled clinic and medication pickup visits as planned; 2) cyclical (attended ≥1 visits late >28 days but returned to and remained in care); or 3) disengaged (missed a scheduled visit by >28 days and had no evidence of return).

**Findings:** We enrolled 159,429 adult participants (61% female, median age 33). Of the 513,322 interactions observed in the 12 months after initiation, 53% occurred as planned, 22% were late ≤28 days late, 9% were >28 days late, and 17% were scheduled but never attended. In 0-6 months after initiation, 51% clients were continuously engaged, 12% cyclically engaged, and 33% disengaged. Two thirds of disengagers (21% of cohort) did not return after the initiation visit. During months 7-12, most clients who had been continuously engaged in months 0-6 (54%) remained continuous, while 18% moved to cyclical engagement. Among cyclical engagers in months 0-6, nearly half (47%) moved to being continuously engaged by month 12. Only 34% of the study population remained engaged continuously by the end of the 12-month period.

**Interpretation:** Fewer than 60% of clients initiating ART care between 2018 and 2022 at Zambian facilities remained continuously engaged at month 6 and 34% at month 12. Cyclical engagement and frequent interruptions should be accepted as the norm and models of service delivery designed to accommodate them.

**Funding:** Funding for this study was provided by the Gates Foundation under INV-031690 to Boston University (SR principal investigator and award recipient). www.gatesfoundation.org. The funders had no role in study design, data collection and analysis, decision to publish, or preparation of the manuscript. Support for collection of the data used in this study was provided by the U.S. National Institutes of Health’s National Institute of Allergy and Infectious Diseases (NIAID), the Eunice Kennedy Shriver National Institute of Child Health and Human Development (NICHD), the National Cancer Institute (NCI), the National Institute on Drug Abuse (NIDA), the National Heart, Lung, and Blood Institute (NHLBI), the National Institute on Alcohol Abuse and Alcoholism (NIAAA), the National Institute of Diabetes and Digestive and Kidney Diseases (NIDDK) and the Fogarty International Center (FIC) under Award Number U01AI069924. The content is solely the responsibility of the authors and does not necessarily represent the official views of the National Institutes of Health.

## Introduction

Despite the tremendous success of HIV treatment programs in sub-Saharan Africa, treatment interruptions and disengagement from lifelong antiretroviral therapy (ART) remain major challenges to achieving global and national targets^1^. The early treatment period--typically defined as the first 6 months after ART initiation or re-initiation--has long been the interval of highest risk for treatment interruption and disengagement^2–4^. In Zambia, for example, the rate of loss to follow up in the first six months on ART was four times greater than in the ensuing 3.5 years in one early study^5^; for the four studies from Zambia included in a systematic review of retention, it was estimated that 90% of attrition in the first year after initiation occurred in the first six months^6^.

The large body of published literature on retention in ART programs^6–9^ has traditionally focused on long-term outcomes, regarding the first six months after initiation--or even the first 12 months--as a single block of time. It has not examined what happens to ART clients within this period. These aggregate reports conceal client behavior and patterns of care-seeking that are critical to understanding high rates of disengagement during this period, predicting long-term outcomes, and identifying the characteristics of clients who can safely be transferred to lower-intensity models of care even sooner than guidelines currently allow^10^. We recently published data showing that in South Africa, like Zambia a high-HIV-prevalence country in southern Africa, 41% of initiating clients experienced an interruption of >28 days during their first six months on treatment, and only 45% remained continuously in care, with no interruptions, by the 12-month point^11^. Here we report a parallel analysis from Zambia, using retrospective medical record data from more than 500 healthcare facilities to analyze patterns of care between ART initiation, 6 months after initiation, and 12 months after initiation; timing of interruptions and disengagement; and the resource utilization associated with different patterns of care.

## Methods

### Study population

For this analysis, we used anonymized, retrospective, client-level data from Zambia’s electronic medical record system, known as SmartCare, that were provided to the International Epidemiologic Databases for the Evaluation of AIDS (IeDEA)^12^ by the Centre for Infectious Disease Research in Zambia (CIDRZ). The dataset included 543 public sector facilities receiving technical and material support from CIDRZ, comprising roughly 17% of public healthcare facilities in the country. The specific facilities included in the data set changed over time due to changes in Smartcare affecting data collection and data reporting. Overall, facilities in the dataset represented 74% (n=173) of all public healthcare facilities in Lusaka Province and 50% (n=163) in Western Province; facility inclusion in other provinces ranged from 3% to 16%. A breakdown of the facilities included in the study by province and district is available in Supplementary File 1 and illustrated in Supplementary Figure 1. Of these facilities, 69% were urban and 31% rural; 87% were health centers (primary healthcare clinics), while 3% were district hospitals and 10% were hospitals offering a higher level of care.

Inclusion criteria for the analytic sample were 1) observed ART initiation visit on or after 1 January 2018; and 2) ≥ 14 months of potential follow-up reflected in the dataset prior to the censor date for outcome variables. The 14-month endpoint provided a two-month window to observe visit attendance occurring after our 12-month endpoint so that we could correctly classify visits scheduled at 12 months. Records from clients aged <18 or >85 years at date of ART initiation and records with a recorded ART initiation date but no observed visit dates in the datasets—including no visit on reported date of ART initiation—were excluded. The dataset allowed us to observe visit patterns for those who had previously initiated treatment at a facility in the dataset, interrupted treatment, and then returned to another facility in the dataset after a period of disengagement. It did not, however, allow us to observe re-engagement in care at any facilities outside the dataset. Clients who disengaged from care at their initiating facility within our dataset and then re-started treatment at a different facility outside of our dataset appeared as permanent disengagers.

### Data

For each individual in the dataset, we received facility name and location, client sex and age at ART initiation, date of ART initiation, observed dates of all types of clinic visits, date of next scheduled clinic visit, laboratory tests (types and dates) and observed date of death or transfer to another facility if reported. Due to known data concerns around the completeness of capture of visits in the database, we first considered every service interaction logged (e.g. visit, dispensing or laboratory test) as a separate encounter. We then grouped together encounters that occurred within 7 days of each other and defined each 7-day cluster of encounters as a single “event”. The date of the next scheduled event is the latest date in the dataset that the client is expected to return to the facility. This is most commonly the date a client is expected to run out of medications already dispensed, assuming the client has adhered to the expected daily dosing frequency, not interrupted treatment, and not obtained antiretroviral medications from any other source. For example, a “next scheduled event” date may be three months after the last observed actual visit date; this assumes that the client received a three-month supply of medications and is due back the day they are expected to run out.

The earliest observation in the analytic data set was 1 January 2018. The dataset was censored on 28 February 2023.

### Variables and outcomes

We used the observed and scheduled dates for each event to classify them based on whether they were attended 1) as planned (on schedule); 2) late ≤28 days; 3) late >28 days or 4) a scheduled visit for which no event was observed. Full definitions of visit classifications have been described elsewhere^11^ and are summarized in Table 1A.

**Table 1.** Definitions of visit categories and patterns of engagement.

| (1A) Visit categories |  |  |
| --- | --- | --- |
| Visit category | Definition |  |
| Initiation visit | Visit on observed date of ART initiation. |  |
| Attended as planned | Visits attended on or before the next scheduled date. Where multiple services observed on the same date had a different next scheduled date, we used the later date between the options. |  |
| Attended late ≤ 28 days | Visits attended after but within 28 days of the scheduled date. |  |
| Attended late ≥ 28 days | Visits attended more than 28 days after the scheduled date. |  |
| Scheduled visit not attended | Date at which disengagement was recognized after a scheduled visit was missed. This date is 28 days after the last scheduled, unattended visit. |  |
| (1B) Patterns of engagement |  |  |
| Label | Pattern of engagement observed during months 0-6 after ART | Pattern of engagement observed during months 7-12 after ART |

|  | **initiation** | **initiation** |
| --- | --- | --- |
| Continuous | No visit > 28 days late in first 6 months and next visit scheduled >6 months after initiation (i.e. all visits as planned or late ≤28 days) | Continuous or cyclical at 6 months AND no visit > 28 days late in months 7-12 and next visit scheduled >12 months after initiation (i.e., all visits attended as planned or late ≤28 days) |
| Cyclical | Attended at least 1 visit late by more than 28 days between initiation and month 6 but subsequently re-engaged in care by 6 months after initiation. | Continuous or cyclical at 6 months; attended at least one visit late by more than 28 days between months 7 and 12 but subsequently re-engaged in care by 12 months after initiation. |
| Immediate disengagement | No visits after date of ART initiation; not observed during months 0-6 after ART initiation | Not reported for months 7-12; aggregated under "Disengaged months 0-6" |
| Early disengagement | ≥1 visit after date of ART initiation but last visit ≤ 3 months after initiation | Not reported for months 7-12; aggregated under "Disengaged months 0-6" |
| Late disengagement | ≥1 visit after date of ART initiation but last observed visit occurs 4-6 months after initiation (i.e. no further scheduled or observed visits in 7-12 month period) | Not reported for months 7-12; aggregated under "Disengaged months 0-6" |
| Disengaged months 0-6 | Composite pattern including all those who disengaged in the first 6 months; no visits observed after 6 months | Classified as immediate, early, or late disengagers in first 6 months AND no visits observed during months 7-12 |
| Disengaged months 7-12 | Not reported for months 0-6 | Classified as continuous or cyclical at 6 months; at least 1 visit observed in months 7-12 but ≥1 scheduled visit late by >28 days with no evidence of return during months 7-14 |
| Transferred | Documented transfer to another healthcare facility during month 0-6 | Documented transfer to another healthcare facility at any time from 0-12 months |
| Died | Death recorded at any time from 0-6 months | Death recorded at any time from 0-12 months |

We used the visit categories in Table 1A to define engagement patterns for months 0-6 and months 7-12 after initiation or re-initiation (Table 1B). “Continuous engagement” required a client to attend all visits as planned or within 28 days of schedule throughout the observation period and have a scheduled or observed visit at the outcome end point of day 183 for months 0-6 and day 365 for months 7-12. Clients who attended some visits late >28 days but returned to care before the measured endpoint (6 or 12 months) were deemed “cyclical.” We considered clients as disengaged from care if they missed a scheduled visit by >28 days and had no evidence of return for clinic visit or medication dispensing occasion during the remaining observation time.

As shown in Table 1B, disengagement was further classified by its timing. For the 0-6 month period, those who initiated but were never observed to return for their first follow-up visit after initiation were deemed as “immediate” disengagers. Those who had at least one return visit but disengaged within 90 days of ART initiation were labeled “early” disengagers. Those who disengaged after 90 days but before 183 days were defined as “late” disengagers. For the 7-12 month period, participants who had already disengaged during months 0-6 were called “disengaged months 0-6” and all those who disengaged after day 183 “disengaged months 7-12”. Participants who attended visits throughout the observation period and had a next scheduled visit date after the 12-month endpoint were deemed engaged in care at 12 months.

### Statistical analysis

We first described frequencies and proportions of participant characteristics at ART initiation stratified by year of initiation and engagement pattern as defined at 6- and 12-months post ART initiation. We also synthesized these findings in an alluvial chart, which visualizes the proportion of participants in each engagement pattern by 6-month period and allowed us to see shifts between engagement types over the two time periods.

Next, we considered predictors of disengagement at our two outcome points, reporting crude risk ratios with 95% confidence intervals. We adjusted each crude estimate for other predictors using log-linear regression models (aRR). The predictors used were the same across the two time periods, with participants’ engagement pattern during months 0-6 also being considered as a predictor of disengagement during months 7-12.

We then quantified resource utilization. We calculated and reported utilization in four resource categories: 1) number of visits and dispensing occasions; 2) ART dispensing quantities; 3) numbers of CD4 counts and HIV viral load tests performed; and 4) number of non-HIV laboratory tests and medications utilized. We report resource utilization by 12-month engagement pattern and by year of ART initiation. Dispensing quantities were estimated using EMR fields for dispensing start and end dates. Medications are typically dispensed in multiples of 30 (since tablets come in bottles of 30, 60, 90, or 180), but the date fields in the EMR occasionally resulted in non-standard dispensing quantities for the analysis. Finally, to detect trends in numbers of visits over time, we created line graphs plotting the median number of visits by pattern of engagement per year.

### Ethics statement

Analysis of the de-identified datasets was approved by the University of the Witwatersrand Human Research Ethics Committee (Medical) under protocol M1902105 and by the Boston University Medical Center IRB under protocol H-38815 for the use of data with a waiver of consent. The dataset for this analysis and permission for the analysis were obtained from the IeDEA consortium. The IeDEA-SA Collaboration has ethics approval from the Human Research Ethics Committees of the University of Cape Town (084/2006). Ethics approval was also received from the University of Zambia Biomedical Research Ethics Committee (UNZA-BREC) (IRB00001131).

## Results

### Study population

We enrolled a total of 159,429 individual participants who initiated ART between 1 January 2018 and 31 December 2021 (Table 2). The sex and age of participants remained constant over time, but the cohort became slightly less urban, with 83% of participants in cities or peri-urban areas in 2018 and 75% in 2021, and increasingly dominated by sites in Lusaka Province. We also observed changes in the ART regimen provided at ART initiation, with first-line 3TC-TDF-DTG increasing from only 3% of participants who initiated in 2018 to 92% of those who initiated in 2021. First-line use of 3TC-TDF-EFV decreased over the same period from 59% to 1% and first-line FTC-TDF-EFV from 33% to 6%.

**Table 2.** Baseline characteristics of population, by year of treatment initiation.

| Characteristic | 2018 | 2019 | 2020 | 2021 | Total |
| --- | --- | --- | --- | --- | --- |
| N | 56,604 | 42,378 | 37,288 | 23,159 | 159,429 |
| Sex (self-reported) |  |  |  |  |  |
| Female | 35,550<br>(63%) | 24,934<br>(59%) | 22,730<br>(61%) | 14,455<br>(62%) | 97,669 (61%) |
| Male | 21,054<br>(37%) | 17,444<br>(41%) | 14,558<br>(39%) | 8,704 (38%) | 61,760 (39%) |
| Age at ART initiation<br>(median [IQR]) | 33[27, 40] | 34 [27, 41] | 33 [27, 40] | 33 [27, 40] | 33 [27, 40] |
| Setting |  |  |  |  |  |
| Urban | 46,765<br>(83%) | 34,356<br>(81%) | 29,427<br>(79%) | 17,337<br>(75%) | 127,885<br>(80%) |
| Rural | 2,322 (4%) | 2,890 (7%) | 3,045 (8%) | 2,077 (9%) | 10,334 (6%) |
| Unknown | 7,517 (13%) | 5,132 (12%) | 4,816 (13%) | 3,745 (16%) | 21,210 (13%) |
| Province |  |  |  |  |  |
| Lusaka | 37,330<br>(66%) | 30,549<br>(72%) | 28,716<br>(77%) | 19,515<br>(84%) | 116,110<br>(73%) |
| Southern | 2,422 (4%) | 561 (1%) | 302 (1%) | 185 (1%) | 3,470 (2%) |
| Western | 5,528 (10%) | 6,398 (15%) | 3,885 (10%) | 2 (0%) | 15,813 (10%) |
| Other | 11,324<br>(20%) | 4,870 (11%) | 4,385 (12%) | 3,457 (15%) | 24,036 (15%) |
| ART regimen at initiation |  |  |  |  |  |
| 3TC-TDF-DTG | 1,690 (3%) | 19,578<br>(46%) | 26,750<br>(72%) | 21,389<br>(92%) | 69,407 (44%) |
| 3TC-TDF-EFV | 33,329<br>(59%) | 14,956<br>(35%) | 5,942 (16%) | 295 (1%) | 54,522 (34%) |
| FTC-TDF-EFV | 18,837<br>(33%) | 6,313 (15%) | 1,592 (4%) | 84 (0%) | 26,826 (17%) |
| Other | 2,748 (5%) | 1,531 (4%) | 3,004 (8%) | 1,391 (6%) | 8,674 (5%) |

### Engagement patterns in the first 6 and 12 months on ART

We found that in the first 6 months after treatment initiation, just over half of participants (51%) were engaged in care continuously, 12% were engaged cyclically, and 30% had disengaged from care (Table 3 and Figure 1). A substantial proportion of those who initiated care (21%) were never observed returning to the facility again within our dataset (immediate disengagement), while an additional 3% disengaged from care within 90 days of treatment initiation (early disengagement) and 8% in months 4-6 (late disengagement). Just under 4% of those who initiated ART had a record of transfer to a facility outside of our dataset, and 1.1% died during the early treatment period. Patterns of engagement evolved over the period of observation, with fewer cyclical engagers and more immediate disengagers in 2021 than in 2018.

**Figure 1.**
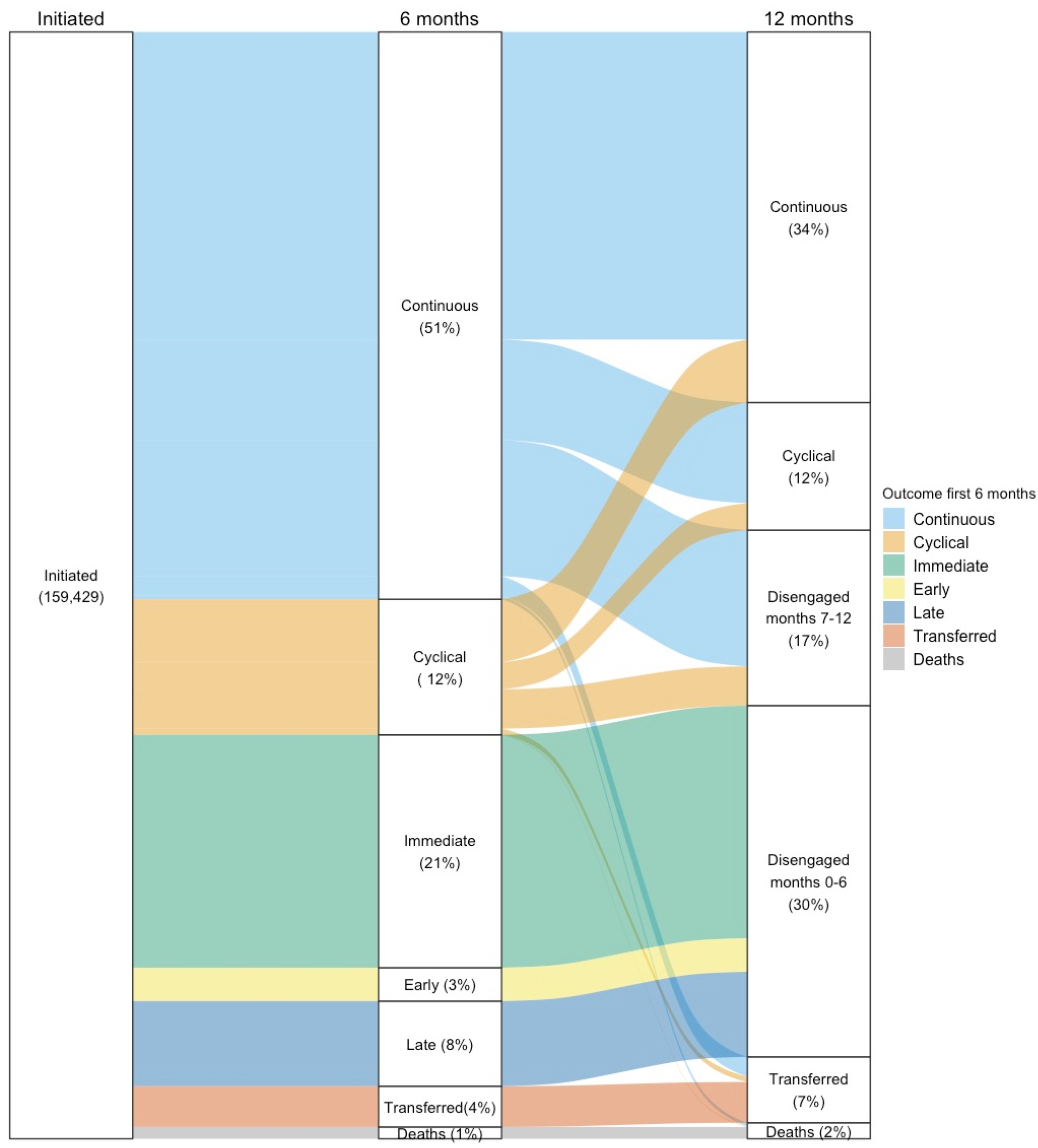
Alluvial chart of patterns of engagement in the first year after treatment initiation.

**Table 3.** Patterns of engagement in months 0-6, by year of initiation.

| Engagement pattern | Year of initiation |
| --- | --- |

| months 0-6 | 2018 | 2019 | 2020 | 2021 | Total |
| --- | --- | --- | --- | --- | --- |
| N | 56,604 | 42,378 | 37,288 | 23,159 | 159,429 |
| Continuous | 24,503 (43%) | 24,091 (57%) | 21,092 (57%) | 12,021 (52%) | 81,707 (51%) |
| Cyclical | 9,171 (16%) | 6,009 (14%) | 2,988 (8%) | 1,373 (6%) | 19,541 (12%) |
| Immediate | 12,888 (23%) | 6,729 (16%) | 7,859 (21%) | 6,052 (26%) | 33,528 (21%) |
| Early | 3,084 (5%) | 1,062 (3%) | 421 (1%) | 246 (1%) | 4,813 (3%) |
| Late | 5,425 (10%) | 2,670 (6%) | 2,438 (7%) | 1,731 (8%) | 12,264 (8%) |
| Transferred | 1,164 (2%) | 1,461 (3%) | 1,986 (5%) | 1,268 (6%) | 5,879 (4%) |
| Died | 369 (0.7%) | 356 (0.8%) | 504 (1.4%) | 468 (2.0%) | 1,697 (1.1%) |

Among those defined as continuously in care in months 0-6, most experienced very short treatment pauses--occasions of being 1-28 days late to the next scheduled clinic visit or medication pickup. Sixty-one percent of continuous engagers were late by less than one week, 32% were late by more than one but less than two weeks, and 18% were late by 2 to 4 weeks at least once during this period. Over the course of follow up, very few participants in the cohort--only 1% (1,679/159,429)--visited a facility within our dataset other than the facility at which they initiated.

Just over half (54%) of the 34% of participants who had a continuous pattern of engagement during months 0-6 remained continuous during months 7-12 (Figure 2, Table 4). Nearly a fifth (18%) of those previously continuous, though, shifted to a pattern of cyclical engagement in their second half-year period. More surprisingly, among those who had engaged in a cyclical pattern during months 0-6, nearly half (46%) became continuous in months 7-12. A large minority (29%) disengaged during the second period; only 20% remained cyclical in both periods.

**Table 4.** Patterns of engagement in months 7-12 by engagement pattern in months 0-6.

| Engagement pattern months 0-6 | N months 0-6 | Engagement pattern months 7-12 |  |  |  |  |
| --- | --- | --- | --- | --- | --- | --- |
|  |  | Continuous | Cyclical | Disengaged in months 7-12 | Transferred | Died |
| N | 101,248 | 53,380 | 18,366 | 25,276 | 3,633 | 604 |
| Continuous | 81,707 | 44,297 (54%) | 14,463 (18%) | 19,612 (24%) | 2,823 (3%) | 512 (1%) |
| Cyclical | 19,541 | 9,083 (46%) | 3,903 (20%) | 5,664 (29%) | 799 (4%) | 92 (0%) |

**Figure 2.**
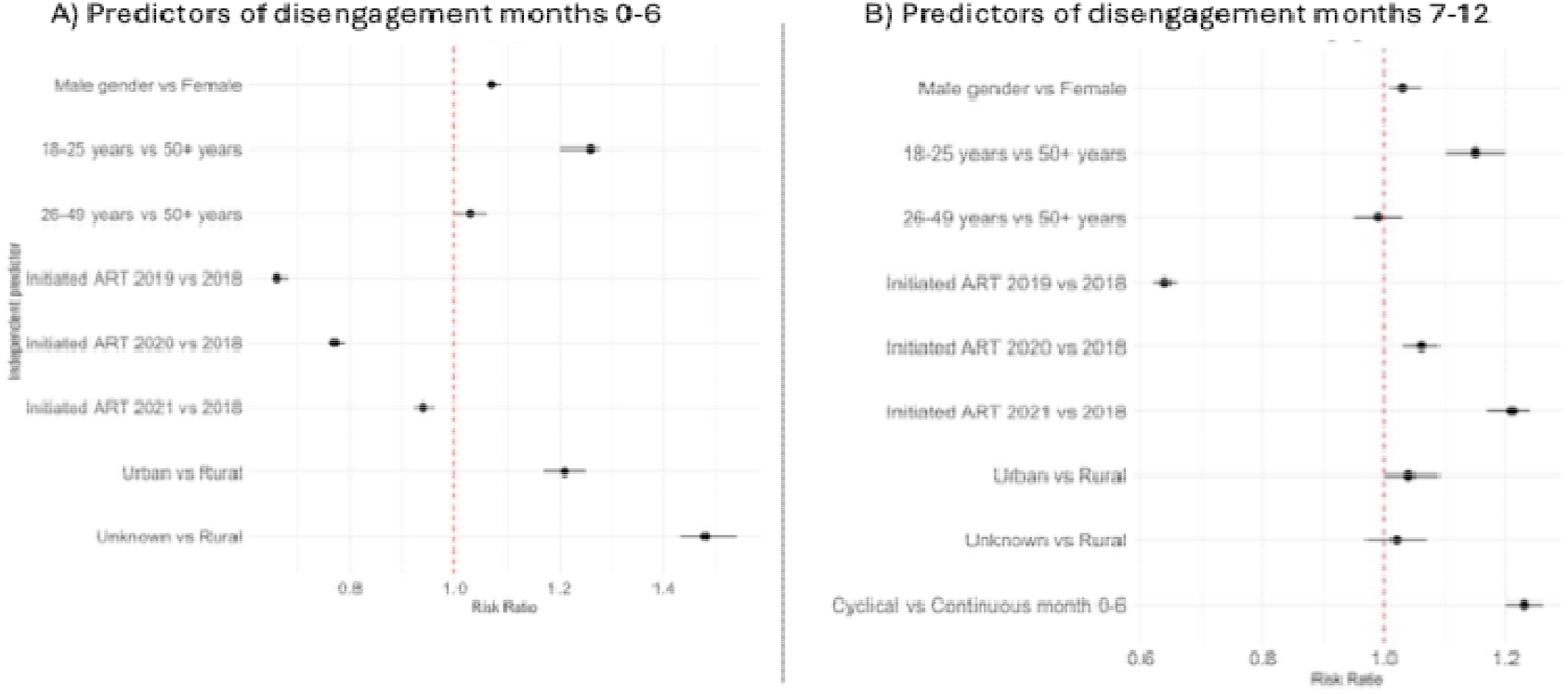
Predictors of disengagement from care in (A) months 0-6 and (B) months 7-12*. *(B) includes only participants who were continuous or cyclical (i.e. still in care) at the end of month 6

Overall, only 28% of the sample remained continuously in care, with no interruptions >28 days, over the course of both periods (month 0 to month 12). Fifteen percent of the cohort experienced at least one interruption of >28 days during the 12-month period, while remaining in care. Total disengagement at the end of 12 months (including those who disengaged in months 0-6) was 47%. When we stratified results by sex (Supplementary Table 2), we found that men were more likely to die during both the first and second 6-month intervals after treatment initiation, but other patterns of engagement were similar between the sexes.

### Predictors of disengagement

We illustrate crude and adjusted relative risks of disengagement at 6 months and 12 months in Figure 2 and Supplementary Table 3. Men had a very slightly higher risk of disengagement than women in both time periods (adjusted relative risk (ARR) 1.07; 95% CI 1.06-1.09 after 6 months and 1.03 (1.01-1.06) after 12 months). Those aged 18-25 had 1.26 (1.23-1.30) times the ARR of being disengaged at the end of the first 6 months than did older age groups, while those aged 26-49 years had similar risk to those aged over 50 (1.03; 1.00-1.06). The effect of age was smaller in months 7-12 but still statistically significant. Disengagement in months 0-6 was also more likely among those residing in urban areas than in rural areas (1.21; 1.17-1.25).

Year of initiation had an inconsistent association with risk of disengagement: disengagement risk was lower in the three years following 2018 than in 2018 during the first six-month period (though gradually increasing over that period) but rose enough in the second six-month period that the ARR of disengagement in months 7-12 in 2021 compared to 2018 was 1.21 (1.17-1.24).

Finally, as might be anticipated, compared to participants who maintained continuous engagement during the first six months, participants with cyclical engagement in the first period were at higher risk of disengagement in the second period (1.23, 1.20-1.26).

### Visit timing

A total of 513,322 events were observed to occur or scheduled to occur in the 12 months following initiation for our analytic cohort (Table 5). More than half (53%) of these were observed as planned (on time), 22% were late by less than 28 days, 9% were late by more than 28 days, and 17% were scheduled to take place by the 12-month endpoint but were never observed in our 14-month observation period. Among those classified as continuously engaged in care at 12 months, 67% of all events in months 0-12 were observed as planned, 27% were late less than 28 days, and 6% were late more than 28 days, with the latter all among participants who engaged cyclically during months 0-6 but shifted to a continuous pattern in months 7-12.

**Table 5.** Resource utilization in the first year after treatment initiation by engagement pattern in months 7-12.

| Resource | Engagement pattern months 7-12 |  |  |  |  |  | Total |
| --- | --- | --- | --- | --- | --- | --- | --- |
|  | Continuous | Cyclical | Disengaged Month 0-6 | Disengaged Month 7-12 | Transferred | Died |  |
| N | 53,416<br>(33.5%) | 18,366<br>(11.5%) | 46,960<br>(29.5%) | 27,284<br>(17.1%) | 10,997<br>(6.9%) | 2,406<br>(1.5%) | 159,429 |
| Total events after initiation in months 0-12 | 250,023 | 66,859 | 71,471 | 92,566 | 26,409 | 5,994 | 513,322 |
| Event type |  |  |  |  |  |  |  |
| As planned | 167,770<br>(67%) | 32,262<br>(48%) | 17,049<br>(24%) | 39,296<br>(42%) | 10,569<br>(40%) | 2,806<br>(47%) | 269,752<br>(53%) |
| Late <28 days | 67,744<br>(27%) | 14,884<br>(22%) | 5,898<br>(8%) | 17,055<br>(18%) | 4,229<br>(16%) | 804<br>(13%) | 110,614<br>(22%) |
| Late >28 days | 14,333<br>(6%) | 19,706<br>(29%) | 1,564<br>(2%) | 8,931<br>(10%) | 1,732<br>(7%) | 257<br>(4%) | 46,523<br>(9%) |
| Scheduled visit not observed | 176<br>(0%) | 7<br>(0%) | 46,960<br>(66%) | 27,284<br>(29%) | 9,879<br>(37%) | 2,127<br>(35%) | 86,433<br>(17%) |
| Number of events |  |  |  |  |  |  |  |
| Mean (SD) | 6 (2) | 5 (2) | 2 (0.8) | 3 (1) | 3 (2) | 3 (2) | 4 (2) |
| Median [Q1, Q3] | 6 [5, 7] | 5 [4, 7] | 1 [1, 2] | 3 [2, 4] | 2 [1, 3] | 2 [1, 4] | 4 [2, 6] |
| Number of ART dispensing occasions |  |  |  |  |  |  |  |
| Mean (SD) | 6 (2) | 5 (2) | 2 (1) | 3 (2) | 3 (2) | 3 (2) | 4 (3) |
| Median [Q1, Q3] | 6 [5, 7] | 5 [4, 6] | 1 [1, 2] | 3 [2, 4] | 2 [1, 3] | 2 [1, 4] | 3 [2, 6] |
| Number of clinic visits |  |  |  |  |  |  |  |
| Mean (SD) | 7 (2) | 6 (2) | 2 (1) | 4 (2) | 3 (2) | 3 (2) | 4 (3) |
| Median [Q1, Q3] | 6 [5, 8] | 6 [4, 7] | 1 [1, 2] | 3 [2, 5] | 2 [1, 4] | 2 [1, 4] | 4 [2, 6] |
| Other resources used |  |  |  |  |  |  |  |
| Documented CD4 count | 14,164<br>(27%) | 4,364<br>(24%) | 5,518<br>(12%) | 4,597<br>(17%) | 1,408<br>(13%) | 383<br>(16%) | 30,434<br>(19%) |
| Documented viral load test | 35,959<br>(67%) | 10,796<br>(59%) | 2,547<br>(5%) | 6,516<br>(24%) | 1,701<br>(15%) | 414<br>(17%) | 57,933<br>(36%) |
| Documented non-ART labs | 12,871<br>(24%) | 3,882<br>(21%) | 4,866<br>(10%) | 3,881<br>(14%) | 1,278<br>(12%) | 393<br>(16%) | 27,171<br>(17%) |
| Dispensed medication other than ART | 36,496<br>(68%) | 10,523<br>(57%) | 21,028<br>(45%) | 15,433<br>(57%) | 5,856<br>(53%) | 1,369<br>(57%) | 90,705<br>(57%) |

### Resource utilization

Resource utilization is also reported in Table 5. Those continuously engaged in care had the highest resource utilization across all resource categories. They reported a median of 6 (IQR 5-7) total events, which included 6 (5-7) medication dispensing occasions and 4 (2-5) clinic visits over the first year after treatment initiation. Two thirds (67%) of them had a documented viral load test, 27% a CD4 count, and 24% a non-ART laboratory test and 68% were dispensed a medication other than ART at some point in the year.

We further stratified resource utilization by year of initiation and engagement pattern (Figure 3, Supplementary Table 4). For those continuously in care in months 7-12, the median number of events per patient during the first year fell from 7 to 5, while for those with cyclical engagement during this period, the median decreased from 6 to 5. Decreases in events/patient/year over time coincided with increases in ART dispensing duration from a median of 65 days (IQR 53-75) in 2018 to 90 days (75-105) in 2021 among continuous engagers and from 60 days (48-72) to 90 days (70-103) among cyclical engagers.

**Figure 3.**
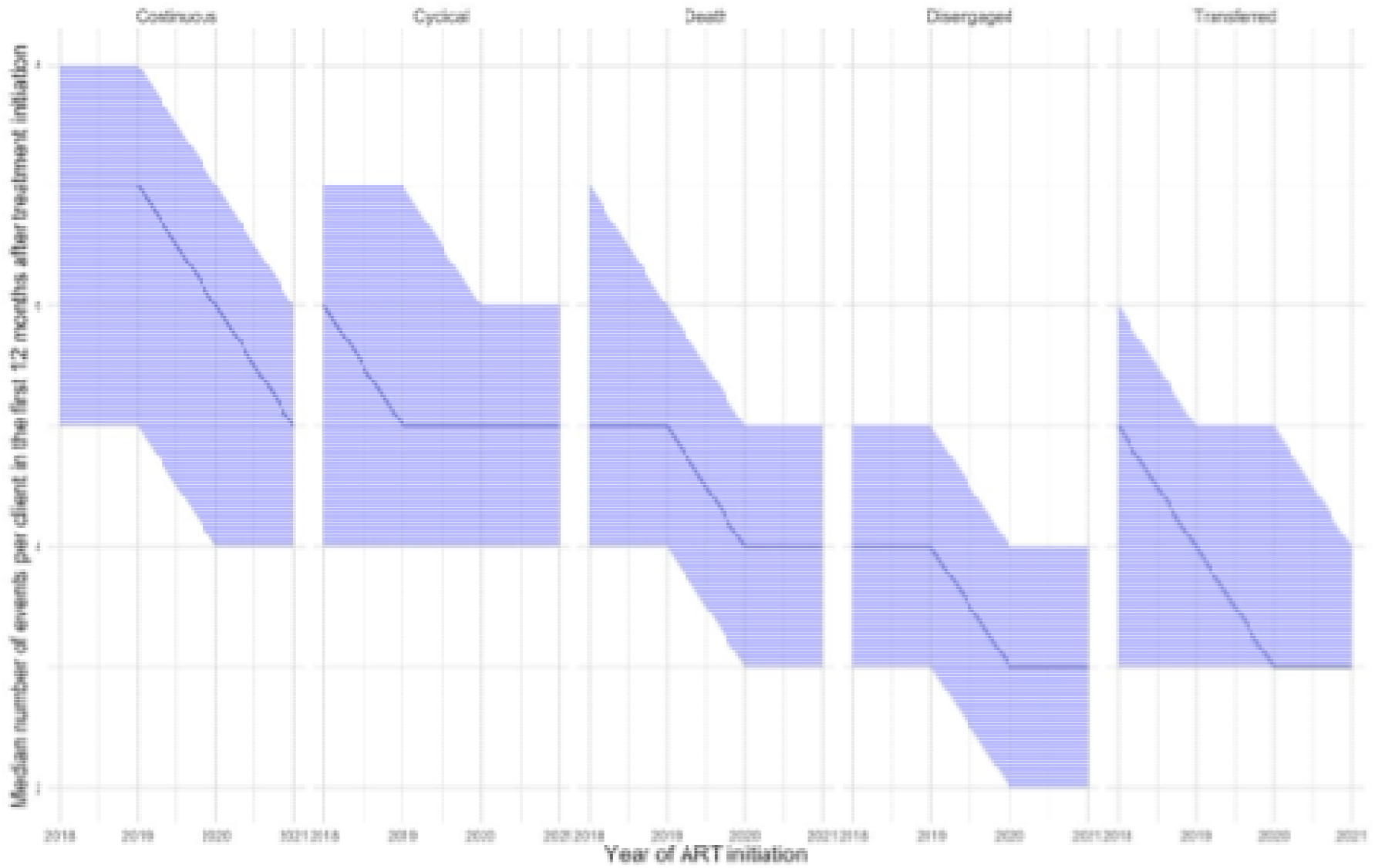
Median number of events per patient in the first 12 months after initiation, by pattern of engagement in months 7-12 and calendar year of initiation*. *Shading indicates interquartile range.

Utilization of other resources (laboratory tests and non-ART medications) generally reflected patients’ patterns of engagement. Only a fifth of those in the cohort (19%) had CD4 counts, a test that is typically done at initiation and then not thereafter. The small proportion of participants with baseline CD4 counts may reflect a shift in Zambia’s guidelines away from CD4 count testing as viral load testing was expanded during the years of the study^13^. Only two thirds of those continuously in care had a documented viral load test result, despite the guidelines calling for this test for all patients at 6 months. Other laboratory tests were relatively rare, with fewer than a quarter of patients having documentation of any other tests even among those continuously in care. More than two thirds of those continuously in care, however, and well over half (57%) of all patients, had records of non-ARV medications being dispensed, primarily isoniazid for tuberculosis prevention and co-trimoxazole for prevention of other infections.

## Discussion

In this analysis of more than 150,000 adults who initiated or re-initiated HIV treatment in Zambia between 2018 and 2021, we found that more than a fifth (21%) did not return to the clinic after the initiation visit. Only 28% remained continuously in care throughout both 6-month periods, with no interruptions >28 days during their first 12 months, and 15% had at least one interruption of 28 days or more over the 12-month period. Nearly half (47%) had disengaged from care by the end of 12 months. Equally important, a cyclical pattern of engagement in the first six-month period predicted disengagement in the second period, suggesting that for some clients, early interruptions are a signal of obstacles to long-term retention.

Based on this study’s findings, it is clear that the archetype patient who initiates ART once, remains in care at the initiating clinic, and never misses a visit or medication refill has become the exception, rather than the rule, for Zambia’s ART program. As was hypothesized several years ago when the phrase “cyclical cascade” was first proposed^14^, both short interruptions and disengagement from care are more common than not. Treatment guidelines and procedures are needed that recognize and accommodate this reality, rather than regarding those who are late for visits or experience disengagement and re-engagement as non-compliant.

Results from this study in Zambia showed both similarities and differences to our prior study in South Africa, which used similar data and methods^11^. General patterns of engagement were similar in both countries, with fewer than half of initiators remaining continuously engaged after 12 months and high frequencies of interruption and disengagement. Age was a major predictor of disengagement in both countries. Somewhat in contrast to results from South Africa, however, we found that in Zambia a cyclical pattern of engagement in the first six-month period did not necessarily augur a negative outcome in the second period. Of patients in our Zambia cohort who had one or more treatment interruptions in the first six-month period, nearly half (46%) had settled into a pattern of continuous engagement by the end of the second period. At the same time, nearly a third (29%) went on to disengage from care in the second period. In South Africa, conversely, while 40% of cyclical engagers from the first period became continuous in the second period, most of the rest (38%) remained cyclical, with only 14% disengaging entirely in the second period.

Unfortunately, we do not know what proportion of clients in Zambia who shifted from cyclical to continuous care between the first and second period benefited from active tracing support, such as phone calls or home visits, to encourage them to return, nor the effectiveness of such interventions. Using earlier data from a similar patient cohort in Zambia, Beres et al (2021) concluded that tracing had no effect on the probability of patients returning to care after disengagement^15^. An earlier study in eastern Africa reported that tracing led to a small (3%) increase in the proportion of patients returning to care^16^. While we cannot estimate how many in our cohort would have returned without intervention, therefore, the high percentage who did return--and the large proportion of continuous engagers who were 1-28 days late for at least one visit--suggests that short interruptions cannot always be regarded as an indication of disengagement or a need for active follow up. At the same time, however, cyclical engagement in the first period did remain a significant risk factor for disengagement in the second period, with an adjusted risk ratio of 1.23. Understanding the characteristics of patients who return voluntarily, compared to those who do not, would help programs target their interventions to those most likely to benefit.

We were surprised to observe only a small difference in engagement patterns between males and females, in view of the prevailing concern about HIV treatment outcomes for men^17,18^. During the first six-month period, 32.2% of men and 31.4% of women disengaged from care; an additional 16.8% and 17.3% disengaged during the second period, respectively. These results are consistent with an earlier (2014-2020) study of adult patients in Zambia’s Southern Province; sex was not a predictor of retention in care in that study^19^. Further research to explain the apparent contradiction between the accepted wisdom that men suffer substantially poorer outcomes than women and the data reported here and elsewhere would be helpful. ^20^

Other predictors of disengagement in our study included younger age, later year of ART initiation, and urban location. Age is consistently a risk factor for poorer outcomes^11,21,22^. Outcomes at 6 months appear to have deteriorated sharply between 2018 and 2019 and then gradually improved in the following years, a pattern that may reflect the initial effect of universal treatment eligibility (“test and treat”), which was adopted in Zambia in 2017^23^, and may also be due in part to changes in data collection procedures and/or to treatment program evolution. Finally, we observed better outcomes for patients at urban clinics, suggesting that remaining continuously in care may be more challenging for those living in rural areas.

The expansion of multi-month dispensing of antiretroviral medications is evident in our study, as the median number of interactions between patients and the healthcare system fell from 7 to 5 per year for those remaining continuously in care. Longer dispensing intervals were not associated with differences in engagement patterns; by the last year of observation, both continuous and cyclical engagers received a median of three months of medications at a time. Of more concern may be the finding that only two thirds of those continuously in care at the end of 12 month had a viral load test result documented in their records; while some missing test results may reflect incomplete data entry, it is likely that many patients did not receive viral load tests at all during their first year.

Our study had a number of limitations. Most important, we were not able to observe unofficial or “silent” transfers to facilities outside our data set, leading to a likely over-estimate of true disengagement. A recent study using data from the same population in Zambia estimated that 27% of patients presenting as ART naive at initiation had suppressed viral loads and were thus likely undisclosed transfers from other facilities^24^. Another, also in this Zambia population, reported high rates of silent transfers, though often after a gap of more than 6 or 12 months^25^, which would have led these individuals to be classified a disengaged in our study regardless of later return. Second, we know that like most observational, routinely collected clinical datasets, the one used in this study was missing some data that may have affected our results, such as unrecorded clinic visits or laboratory tests^26,27^. Third, as is clear from the results presented here, “interruptions” in HIV treatment as measured by clinic visits and dispensing intervals range from a few days to many months. It is likely that some patients with interruptions of a few weeks, or even several months, still have supplies of medications on hand and remain adherent to treatment, despite the gaps in their records. And finally, our study period overlapped with COVID-19 related restrictions in Zambia. Constraints on accessing care may have affected clinic visits reported in our data set, particularly in 2020 and 2021. An updated analysis, incorporating utilization of care in 2023 and 2024, would help to confirm post-COVID visit behavior.

## Conclusion

Despite these limitations, our study draws attention to the importance of understanding treatment interruptions and disengagement during the early treatment period. We demonstrate the frequency of both short and long interruptions, even for ART clients who remain in care, and the associated risk of early interruptions with longer-term disengagement. The needs of continuous and cyclical engagers may differ from those who disengage from care during the first 6 months after initiation. Determining what happens to the fifth of all initiators who are immediate disengagers--those who start ART but then never return to the same clinic--is a priority; if most of them self-transfer to another facility, the challenge facing Zambia’s HIV program is completely different than if most of them actually disengage immediately after their initiation visit. Understanding the timing of interruptions and patterns of re-engagement in care is also critical to improving the efficiency of interventions to improve retention in care, such as active tracing of those who miss visits. We conclude treatment guidelines and procedures need to accommodate the reality that short and long interruptions and cyclical engagement are more common than not, rather than treating those who are late for visits or experience disengagement and re-engagement as non-compliant.

## Data Availability

All of the data used in this manuscript were provided by the IeDEA Southern Africa collaboration. Access can be requested by contacting Dr. Morna Cornell, Project Manager IeDEA-SA.

## Supplementary material

Supplementary Table 1. Facilities included in the analysis, by district

Supplementary Table 2. Patterns of engagement stratified by sex

Supplementary Table 3. Predictors of disengagement in months 0-6 and months 7-12

Supplementary Table 4. Resource utilization in the first year after treatment initiation for those who are engaged in care at 12 months

Supplementary Figure 1. Location of facilities included in the analysis

## Acknowledgements

We wish to acknowledge all study participants who contributed data for this analysis. We also thank the CIDRZ and IeDEA-SA data managers for collating these data.

## Authors’ roles

MB conceived of the study, performed the data analysis, and drafted the manuscript. MM conceived of the study and contributed to the data analysis and interpretation of results. PC contributed to the data analysis. MWM contributed to interpretation of results. TS contributed to interpretation of results. BN contributed to the data analysis and interpretation of results. CBM contributed to interpretation of results. LM contributed to interpretation of results. SS contributed to interpretation of results. KZ contributed to interpretation of results. SR conceived of the study, interpreted the results, and drafted the manuscript. All authors reviewed the manuscript and approved the final version.

## Competing interests

The authors have no competing interests to report. LM, SS, and KZ are representatives of the government agency that is responsible for supervision of the study sites.

## Funding

Funding for this study was provided by the Gates Foundation under INV-031690 to Boston University (SR principal investigator and award recipient). www.gatesfoundation.org. The funders had no role in study design, data collection and analysis, decision to publish, or preparation of the manuscript.

Research reported in this publication was supported by the U.S. National Institutes of Health’s National Institute of Allergy and Infectious Diseases (NIAID), the Eunice Kennedy Shriver National Institute of Child Health and Human Development (NICHD), the National Cancer Institute (NCI), the National Institute on Drug Abuse (NIDA), the National Heart, Lung, and Blood Institute (NHLBI), the National Institute on Alcohol Abuse and Alcoholism (NIAAA), the National Institute of Diabetes and Digestive and Kidney Diseases (NIDDK) and the Fogarty International Center (FIC) under Award Number U01AI069924. The content is solely the responsibility of the authors and does not necessarily represent the official views of the National Institutes of Health.

## Prior presentation

Some of the results in this manuscript were presented previously at the International AIDS Society 2024 conference (AIDS 2024) and are posted in the conference archive.

## Notes

### Competing Interest Statement

The authors have declared no competing interest.

