## Supplementary materials for "Patterns of engagement in care during clients’ first 12 months after HIV treatment initiation in Zambia: a retrospective cohort analysis using routinely collected data"

### Supplementary material

Supplementary Table 1. Facilities included in the analysis, by district

Supplementary Table 2. Patterns of engagement stratified by sex

Supplementary Table 3. Predictors of disengagement in months 0-6 and months 7-12

Supplementary Table 4. Resource utilization in the first year after treatment initiation for those who are engaged in care at 12 months

Supplementary Figure 1. Location of facilities included in the analysis

**Supplementary Table 1. Facilities included in the analysis, by district***

| Province | District | Number of facilities in dataset |
|---|---|---|
| **Central (11% of all public sector facilities in province)** | | **30** |
| | Chibombo | 4 |
| | Chisamba | 3 |
| | Itezhi-tezhi | 2 |
| | Kabwe | 9 |
| | Kapirimposhi | 2 |
| | Luano | 1 |
| | Mkushi | 1 |
| | Mumbwa | 3 |
| | Serenje | 2 |
| | Shibuyunji | 3 |
| **Copperbelt (16% of all public sector facilities in province)** | | **35** |
| | Chingola | 2 |
| | Kalulushi | 2 |
| | Kitwe | 15 |
| | Luanshya | 5 |
| | Mpongwe | 1 |
| | Mufurila | 1 |
| | Ndola | 9 |
| **Eastern (11% of all public sector facilities in province)** | | **40** |
| | Chadiza | 2 |
| | Chipata | 15 |
| | Katete | 4 |
| | Lundazi | 7 |
| | Mambwe | 1 |
| | Nyimba | 5 |
| | Petauke | 5 |
| | Sinda | 1 |
| **Luapula (3% of all public sector facilities in province)** | | **10** |
| | Chembe | 1 |
| | Chiengi | 1 |
| | Kawambwa | 1 |
| | Mansa | 2 |
| | Nchelenge | 3 |
| | Samfya | 2 |
| **Lusaka (74% of all public sector facilities in province)** | | **173** |
| | Chilanga | 12 |
| | Chongwe | 30 |
| | Kafue | 21 |
| | Luangwa | 16 |
| | Lusaka District | 72 |
| | Rufunsa | 22 |
| **Muchinga (3% of all public sector facilities in province)** | | **5** |
| | Chinsali | 1 |
| | Isoka | 1 |
| | Mpika | 2 |
| | Nakonde | 1 |
| **North Western (3% of all public sector facilities in province)** | | **9** |
| | Kabompo | 1 |
| | Kalumbila | 1 |
| | Solwezi | 7 |
| **Northern (4% of all public sector facilities in province)** | | **11** |
| | Kasama | 3 |
| | Luwingu | 1 |
| | Mbala | 3 |
| | Mpulungu | 1 |
| | Mungwi | 1 |
| | Nsama | 1 |
| | Nsenga Hill | 1 |
| **Southern (16% of all public sector facilities in province)** | | **67** |
| | Chikankata | 1 |
| | Chirundu | 13 |
| | Choma | 6 |
| | Kalomo | 1 |
| | Kazungula | 3 |
| | Livingstone | 16 |

| Province | District | Number of facilities in dataset |
| --- | --- | --- |
| | Mazabuka | 7 |
| | Monze | 5 |
| | Namwala | 4 |
| | Pemba | 1 |
| | Siavonga | 3 |
| | Sinazongwe | 7 |
| **Western (50% of all public sector facilities in province)** | | **163** |
| | Kalabo | 26 |
| | Kaoma | 24 |
| | Luampa | 1 |
| | Lukulu | 22 |
| | Mitete | 5 |
| | Mongu | 7 |
| | Mulobezi | 1 |
| | Mwandi | 1 |
| | Nalolo | 16 |
| | Nkeyema | 9 |
| | Senanga | 19 |
| | Sesheke | 2 |
| | Sikongo | 13 |
| | Sioma | 17 |
| **Total (17% of all public sector facilities nationally)** | | **543** |

*Data on total number of facilities in each province taken from https://mfl.moh.gov.zm/site/index.

**Supplementary Table 2. Patterns of engagement stratified by sex**

| Engagement pattern | Men (N=61,760) | Women (N=97,669) | Overall (N=159,429) |
|---|---|---|---|
| **Engagement in months 0-6** | | | |
| Continuous | 31313 (50.7%) | 50394 (51.6%) | 81707 (51.2%) |
| Cyclical | 7254 (11.7%) | 12287 (12.6%) | 19541 (12.3%) |
| Early | 1968 (3.2%) | 2845 (2.9%) | 4813 (3.0%) |
| Immediate | 13015 (21.1%) | 20513 (21.0%) | 33528 (21.0%) |
| Late | 4910 (8.0%) | 7354 (7.5%) | 12264 (7.7%) |
| Transferred | 2369 (3.8%) | 3510 (3.6%) | 5879 (3.7%) |
| Death | 931 (1.5%) | 766 (0.8%) | 1697 (1.1%) |
| **Engagement months in 7-12** | | | |
| Continuous | 20402 (33.0%) | 33014 (33.8%) | 53416 (33.5%) |
| Cyclical | 6887 (11.2%) | 11479 (11.8%) | 18366 (11.5%) |
| Disengaged months 7-12 | 10366 (16.8%) | 16918 (17.3%) | 27284 (17.1%) |
| Disengaged months 0-6 | 18513 (30.0%) | 28447 (29.1%) | 46960 (29.5%) |
| Transferred | 4264 (6.9%) | 6733 (6.9%) | 10997 (6.9%) |
| Death | 1328 (2.2%) | 1078 (1.1%) | 2406 (1.5%) |

**Supplementary Table 3. Predictors of disengagement in months 0-6 and months 7-12**

| Characteristic | Measure | Disengaged months 0-6 (n=50,605) | | | Disengaged months 7-12* (n=25,276) | | |
|---|---|---|---|---|---|---|---|
| | | N | Crude RR (95% CI) | Adjusted RR (95% CI) | N | Crude RR (95% CI) | Adjusted RR (95% CI) |
| Sex | Female | 30,712 | ref. | ref. | 15,656 | ref. | ref. |
| | Male | 19,893 | 1.02 (1.00-1.04) | 1.07 (1.06-1.09) | 9,620 | 1.00 (0.98-1.02) | 1.03 (1.01-1.06) |
| Age (years) | 18-25 | 10,828 | 1.24 (1.20-1.28) | 1.26 (1.23-1.30) | 4,823 | 1.17 (1.12-1.22) | 1.15 (1.10-1.20) |
| | 26-49 | 35,943 | 1.02 (0.99-1.05) | 1.03 (1.00-1.06) | 18,456 | 0.99 (0.96-1.05) | 0.99 (0.95-1.03) |
| | ≥50 | 3,834 | ref. | ref. | 1,997 | ref. | ref. |
| Year of ART initiation | 2018 | 21,397 | ref. | ref. | 9,215 | ref. | ref. |
| | 2019 | 10,461 | 0.65 (0.64-.067) | 0.66 (0.65-0.68) | 5,125 | 0.62 (0.60-0.64) | 0.64 (0.62-0.66) |
| | 2020 | 10,718 | 0.76 (0.75-0.78) | 0.77 (0.76-0.79) | 6,706 | 1.02 (0.99-1.05) | 1.06 (1.03-1.09) |
| | 2021 | 8,029 | 0.92 (0.90-0.94) | 0.94 (0.92-0.96) | 4,230 | 1.15 (1.12-1.19) | 1.21 (1.17-1.24) |
| Setting | Rural | 2,571 | ref. | ref. | 1,705 | ref. | ref. |
| | Urban | 39,884 | 1.25 (1.21-1.30) | 1.21 (1.17-1.25) | 20,534 | 1.06 (1.01-1.10) | 1.04 (1.00-1.09) |
| | Unknown | 8,150 | 1.54 (1.49-1.60) | 1.48 (1.43-1.54) | 3,3037 | 1.03 (0.98-1.09) | 1.02 (0.97-1.07) |
| 6-month outcome | Continuous | 81,707 | | | 19,612 | ref. | ref. |
| | Cyclical | 19,541 | | | 5,664 | 1.20 (1.17-1.24) | 1.23 (1.2-1.26) |

* Limited to subset of participants who were either continuous or cyclical at 6 months.

**Supplementary Table 4. Resource utilization in the first year after treatment initiation for those who are engaged in care at 12 months**

| Resource | Continuous | | | | Cyclical | | | |
|---|---|---|---|---|---|---|---|---|
| | **2018** | **2019** | **2020** | **2021** | **2018** | **2019** | **2020** | **2021** |
| N | 15475 | 17876 | 12825 | 7240 | 8109 | 5765 | 3261 | 1231 |
| Number of events | | | | | | | | |
| Mean (SD) | 6.60 (2.05) | 6.45 (1.84) | 5.53 (1.61) | 5.45 (1.53) | 5.72 (2.09) | 5.34 (1.92) | 4.74 (1.50) | 4.73 (1.56) |
| Median [Q1, Q3] | 7.00 [5, 8] | 7.00 [5, 8} | 6.00 [4, 7] | 5.00 [4, 6] | 6.00 [4, 7] | 5.00 [4, 7] | 5.00 [4, 6] | 5.00 [4, 6] |
| ART dispensing events | | | | | | | | |
| Mean (SD) | 6.46 (2.60) | 6.23 (2.08) | 5.62 (1.99) | 5.31 (1.68) | 5.48 (2.52) | 5.14 (2.16) | 4.84 (1.89) | 4.63 (1.72) |
| Median [Q1, Q3] | 6.00 [5, 8] | 6.00 [5, 7] | 5.00 [4., 6] | 5.00 [4, 6] | 5.00 [4, 7] | 5.00 [4, 6] | 5.00 [4, 6] | 4.00 [3, 6] |
| Number of clinical visits | | | | | | | | |
| Mean (SD) | 7.08 (2.37) | 6.87 (2.06) | 5.94 (1.83) | 5.80 (1.69) | 6.13 (2.37) | 5.70 (2.13) | 5.11 (1.72) | 5.04 (1.73) |
| Median [Q1, Q3] | 7.00 [6, 8] | 7.00 [6, 8] | 6.00 [5, 7] | 6.00 [5, 7] | 6.00 [4, 8] | 5.00 [4, 7] | 5.00 [4, 6] | 5.00 [4, 6] |
| Total number of services provided | | | | | | | | |
| Mean (SD) | 15.9 (7.96) | 14.6 (6.27) | 13.7 (5.68) | 13.2 (5.02) | 13.2 (7.18) | 11.7 (6.02) | 10.9 (4.91) | 10.7 (4.31) |
| Median [Q1, Q3] | 14.0 [10, 20] | 14.0 [10, 18] | 13.0 [9, 17] | 13.0 [9, 17] | 12.0 [8,, 17] | 11.0 [7, 15] | 10.0 [7, 14] | 10.0 [8, 13] |
| Documented CD4 counts | 6655 (43.0%) | 4188 (23.4%) | 2605 (20.3%) | 716 (9.9%) | 2780 (34.3%) | 1056 (18.3%) | 470 (14.4%) | 58 (4.7%) |
| Documented viral load tests | 8772 (56.7%) | 12575 (70.3%) | 9122 (71.1%) | 5490 (75.8%) | 4453 (54.9%) | 3517 (61.0%) | 1959 (60.1%) | 867 (70.4%) |
| Documented non-ART labs | 5608 (36.2%) | 3372 (18.9%) | 2729 (21.3%) | 1162 (16.0%) | 2244 (27.7%) | 941 (16.3%) | 552 (16.9%) | 145 (11.8%) |
| Dispensed medication other than ART | 8002 (51.7%) | 12337 (69.0%) | 9886 (77.1%) | 6271 (86.6%) | 4132 (51.0%) | 3277 (56.8%) | 2167 (66.5%) | 947 (76.9%) |
| Dispensing duration | | | | | | | | |
| Mean (SD) | 65.2 (20.0) | 78.8 (23.3) | 92.8 (27.0) | 92.8 (27.0) | 61.0 (20.4) | 75.7 (24.1) | 89.7 (28.0) | 90.0 (29.2) |
| Median [Q1, Q3] | 64.7 [53, 75] | 75.0 [64, 90] | 90.0 [75, 108] | 90.0 [75, 105] | 60.0 [48, 72] | 72.5 [60, 90] | 87.0 [70, 103] | 90.0 [70, 103] |

**Supplementary figure 1. Map of study sites**

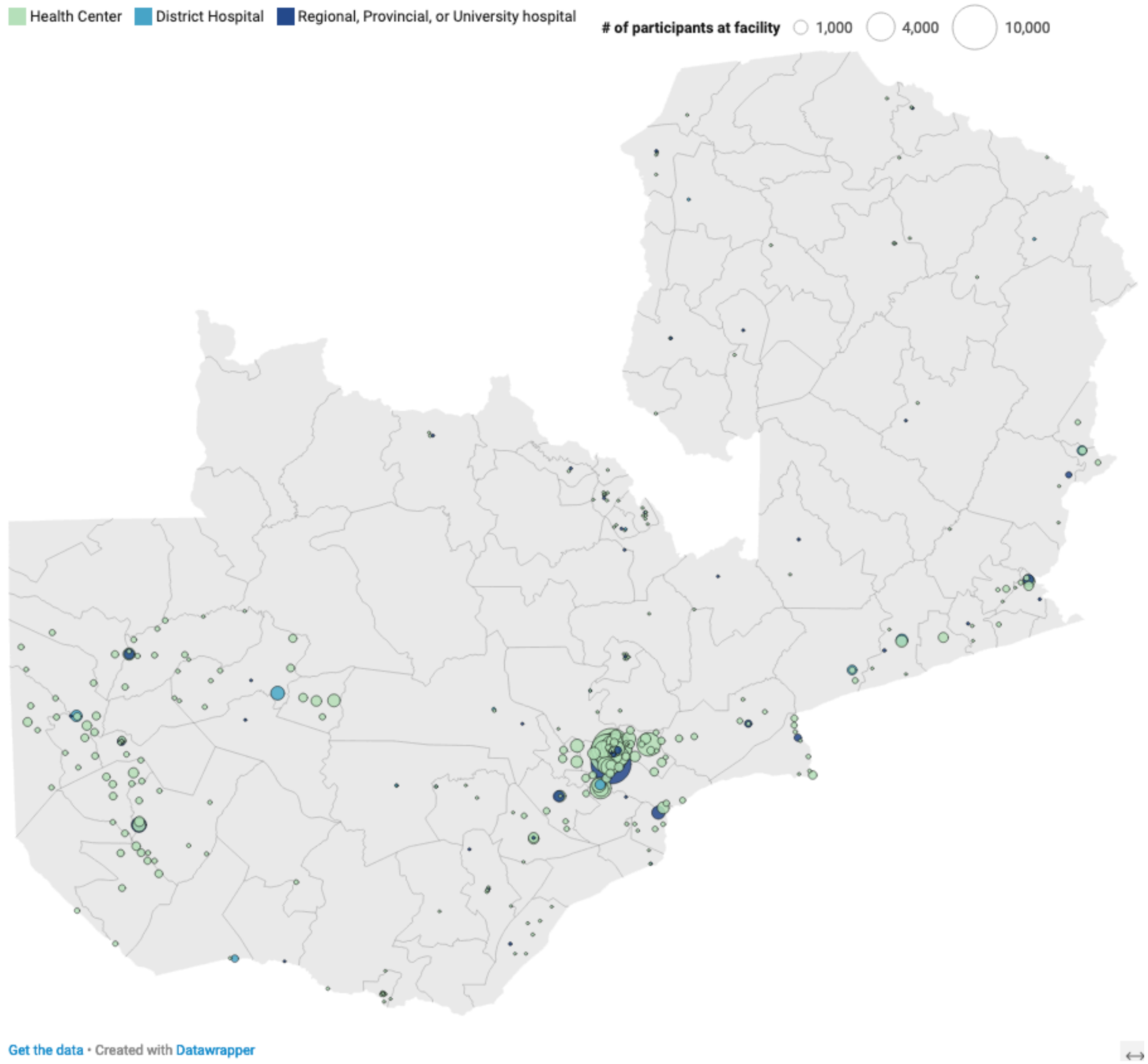
